# Applications of geospatial analyses in health research among homeless people: A systematic scoping review of available evidence

**DOI:** 10.1101/2021.10.26.21265542

**Authors:** Rakibul Ahasan, Md Shaharier Alam, Torit Chakraborty, S M Asger Ali, Tunazzina Binte Alam, Tania Islam, Md Mahbub Hossain

**Affiliations:** Department of Geography, Texas A&M University, College Station, TX 77843 USA; Department of Geography, Florida State University, Tallahassee, FL 32306 USA; Department of Geography, New Mexico State University, Las Cruces, NM 88003 USA; Department of Geosciences, Mississippi State University, Starkville, MS 39759 USA; City Planner, City of Destin, FL 32541 USA; Department of Earth and Environment, Florida International University, Miami FL 33131 USA; Department of Health Promotion and Community Health Sciences, Texas A&M School of Public Health, TX 77843, USA

**Keywords:** geospatial analysis, spatial analysis, GIS, Health GIS, homelessness, systematic review, scoping review

## Abstract

**Background:** The coronavirus pandemic visualized the inequality in the community living standards and how housing is a fundamental requirement to ensure a livable environment. However, even before the pandemic, unequal housing access resulted in more than 150 million homeless people worldwide, and more than 22 million new people were added to this inventory for climate-related issues. This homeless population has a counterproductive effect on the social, psychological integration efforts by the community and exposure to other severe health-related issues.

**Methods:** We systematically identified and reviewed 24 articles which met all three requirements we set forth-i. samples include homeless people, ii. focused on public health-related issues among the same group of people, and iii. used geospatial analysis tools and techniques in conducting the research.

**Result:** Our review findings indicated a major disparity in the geographic distribution of the case study locations-all the articles are from six (6) countries-USA (n = 16), Canada (n = 3), UK (n = 2), and one study each from Brazil, Ireland, and South Africa. Majority of the studies used spatial analysis tools to identify the hotspots, clustering and spatial patterns of patient location and distribution. ArcGIS is the most frequently used GIS application, however, studies also used other statistical applications with spatial analysis capabilities. These studies reported relationship between the location of homeless shelters and substance use, discarded needles, different infectious and non-infectious disease clusters.

**Conclusion:** Although, most studies were restricted in analyzing and visualizing the trends, patterns, and disease clusters, geospatial analyses techniques can be used to assess health problems such as disease distributions and associated factors across communities. Moreover, health and services and accessibility concerns could be well addressed by integrating spatial analysis into homelessness-related research. This may facilitate policymaking for health-issues among the homeless people and address health inequities in this vulnerable population.

## 1. Introduction

The coronavirus pandemic showed the inequality in the living standards among communities and how housing is fundamental to ensure a standard livable environment for all ^1^. Now, more than ever, homelessness needs a balanced understanding of the issue itself and the implementation of long-term solutions. Even before the pandemic, there were more than 150 million homeless people worldwide, about 15 million of which experienced forceful eviction ^2^. An estimated 22 million newly displaced people are added to the current homeless inventory every year due to climate-related events ^2^. However, contrary to popular belief/understanding, homelessness does not only mean street dwellers. It includes housing with inadequate living conditions, living in temporary or emergency accommodation, and other hidden homelessness ^3^. A 2014 study shows that individuals who were once homeless are spread throughout the city, based on the use of homeless services, but had a counterproductive effect on social and psychological integration efforts ^4,5^. Past studies reported the housing prices and location of the highly priced housing options within a community is one of the fundamental reasons of homelessness in any community ^6-8^.

Few of the major reported consequences of being a homeless person alongside the socio-economic vulnerability are substance abuses, addiction, and most significantly, exposure to health issues ^9-12^. Although both the scientific community and practitioners reported struggles in tracking the homeless population and identifying the issues faced by them, recent advancements in geospatial techniques had equipped them to do that. Not many scholarly works were found that explicitly focused on the homeless population-public health issue— geospatial analysis nexus. However, the use of geospatial analysis and techniques in public health issues can be traced back to the 1960s, right after the first reported use of computerized GIS use ^13,14^. Most recent examples of Geographic Information System (GIS) applications in public health emergencies are abundant in the ongoing COVID-19 related studies. Even with the computational efficiency and extensive data requirements, hundreds of these GIS applications related works were published within the first three months of the pandemic ^13,15^.

Researchers from diverse knowledge domains (i.e., Urban Studies, Public Policy, Geography, Public Health, and Economics, etc.) are using the GIS as a useful tool to fight against poverty and homelessness around the world. GIS, as a spatial analysis tool, not only helps in understanding the spatial dynamics of this issue but could be of enormous significance in defining and addressing the issue of affordable housing and homelessness ^16^. So far, the use of GIS in homelessness research is limited only to mapping the locations of the individuals or communities ^17^. However, previous studies had recognized the importance of using geospatial techniques to more accurately visualizing and locating homeless individuals and how they may face health hazards as a result of being homeless. GIS offers much more than mapping and visualization tools to study beyond the locations of the homeless people ^15^. Integrated use of GIS in homelessness studies could provide more insight regarding the causes and consequences of the state. GIS could be used to map and visualize communities with a higher percentage of individuals living on the street (outside of shelter or housing), and communities that make homelessness more likely by integrating spatial analysis in this homelessness research effort. Moreover, by increasing the location-based surveillance Geospatial tools provide a new dimension in homeless research ^18^.

In this study, we aimed to review how past studies utilized GIS and spatial analysis techniques in health research among homeless people. Moreover, we examined the key characteristics of the spatial analysis tools and techniques past studies on homelessness used. We also reviewed the data sources, and variables used in these studies, health-related issues they examined, and issues they encountered in using spatial analysis techniques in their studies. We began with an overview of the characteristics of geospatial technologies used in health research among homeless people and stretched up to exploring opportunities regarding a better integration of geospatial technologies in public health issues, especially among the homeless.

## 2. Methodology

We conducted this scoping review in accordance with the methodological guidance proposed by Arksey and O’Malley. We searched Medline, APA PsycInfo, Academic Search Ultimate, CINAHL (Cumulative Index to Nursing and Allied Health Literature), SocINDEX, Sociology Source Ultimate, Health Source Nursing/Academic Edition, Environment Complete, Social Sciences Full Text, and Health Policy Reference Center using the following search query: (“gis” OR “arcgis” OR “geographic information systems” OR “geographic mapping” OR “spatial analysis” OR “special access” OR “spatial measures” OR “geospatial analysis”) AND (“Homeless” OR “homelessness” OR “unsheltered” OR “unstably sheltered”). This search query was designed to capture homelessness-related literature that mentioned geospatial keywords. Moreover, these keywords were searched in titles, abstracts, subject headings such as MeSH, and general keywords across all fields. No language restriction was applied to the search process. All published studies since the inception of the databases until September 19, 2021, were included during the database searching.

The collection of identified citations was uploaded to Rayyan, a cloud-based systematic review application for two-step screening process. At first two reviewers independently screened all citations based on the following eligibility criteria.

### Inclusion criteria

- **Publication type**- Articles have to be peer-reviewed journal publications.
- **Sample type**- The study sample was current or previously homeless people (irrespective of the variations in working definitions of homelessness in those studies).
- **Study focus**- Investigated any health or health-related problem (determinants of health, health behavior, health outcomes, etc.). In addition to the health-related problems, the articles must utilize any of the geospatial analysis tools and techniques as part of their analysis in investigating health-related outcomes among the homeless population.
- **Availability-** Article full text is available in English.

At the end of the first-round screening, all conflicts were reviewed at the presence of a third reviewer and a consensus was made for each conflict based on discussion. All citations meeting these criteria were selected for full text review. At this stage, two reviewers reviewed the full texts of the articles and determined the eligibility of the articles following the same process as the screening stage.

### Data extraction and synthesis

After the final selection of full texts, a prespecified data extraction was used to extract data from each eligible article. The form included several sections on key variables on study characteristics and health-related indicators, such as the sample characteristics, the location and timeframe of the study, geospatial tools or analyses used in the study, and the key findings of the study. Two reviewers independently extracted data and it was further reviewed at the end of data extraction to address discrepancies based on discussion among the reviewers.

As commonly used in scoping reviews, we conducted a narrative synthesis of the extracted data on key domains emphasizing the review objectives. A tabulation of the key findings alongside a descriptive overview of the literature was prepared. Moreover, visual illustrations of the geographic distributions were prepared to highlight the global scenario of health research on homeless using geospatial approaches.

## 3. Results

### 3.1. Characteristics of the reviewed articles

Even though homelessness is a widely recognized social phenomenon, the number of articles does not reflect the magnitude of the issue. Among 105 articles found from multiple sources, we found 24 articles that met our inclusion criteria (Figure 1). We included these articles in this review. We found a geographical clustering among the case study locations used in these articles. All these articles are from six (6) countries-USA (n=16), Canada (n=3), UK (n=2), and Brazil, Ireland, and South Africa (n=1). All these countries are in the global north (North America and Europe) and have economic efficiency. The absence of studies from the global south is also alarming.

**Figure 1.**
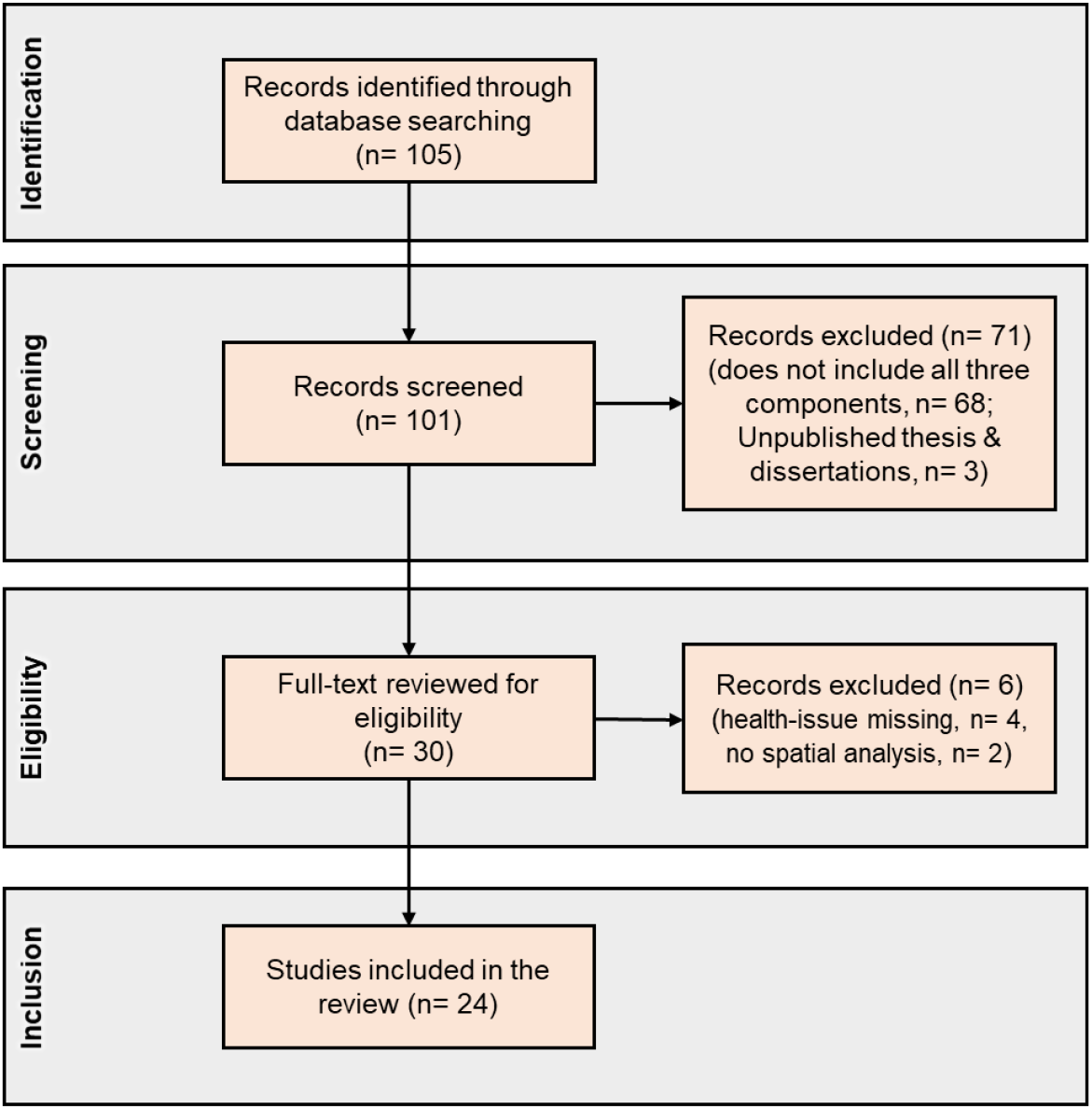
Geospatial analysis in homelessness research literature search and evaluation for inclusion.

From the reviewed articles, there are multiple studies from the same case study site-for example, both Los Angeles and Boston were used multiple times by the same author as a case study location (Citation). The location of these case study locations is shown in figure 2A and 2B. Even though we did not set any earliest year for our review, most of the articles in our review are from the past decade (n= 22). Only 2 of the articles included in this review is from before 2010 (Figure 2C) ^19,20^.

**Figure 2.**
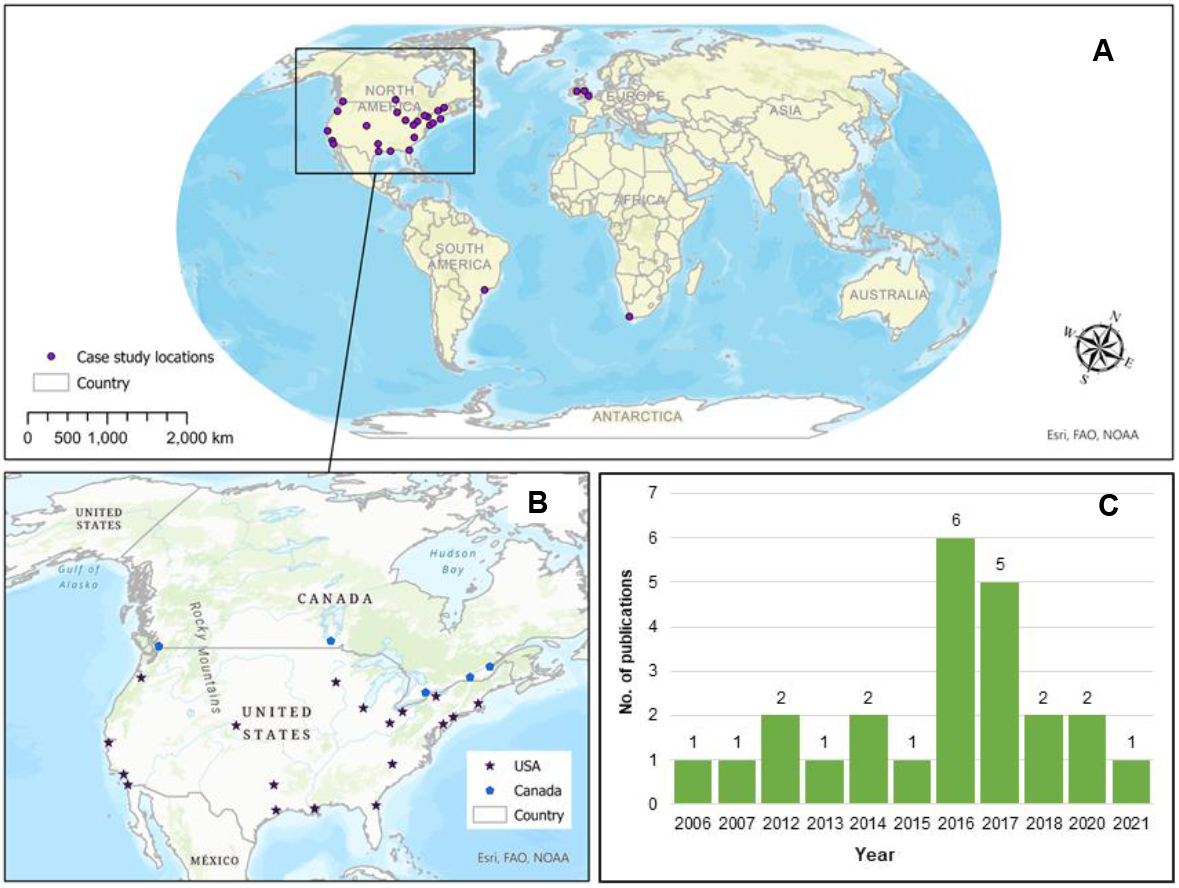
Case study locations and publication trends on health research of the homeless. **A**. All the case study locations from the reviewed studies; **B**. Case study locations from the USA and Canada, which includes 26 out of the 31 case study locations; **C**. Number of publications on homelessness-health issue-GIS nexus

Among the reviewed articles, ArcGIS was the most used GIS platform for geospatial analysis (n = 17). Additionally, several statistical applications were also utilized as spatial analysis tools-Rstudio (n = 3), STATA (n = 3), SAS and Google Earth Engine (n = 1). A majority of these articles used GIS to identify the locations of the homeless people and find out spatial clustering of those of the same group, hotspot analysis, point density analysis, proximity analysis, and mapping and visualization tools were the most frequently used GIS tools. On the other hand, few studies used regression analysis to examine the relationship between the homelessness characteristic and other related variables. Social and demographic variables, race and ethnicity, economic characteristics, geographic location and distribution of homeless people, and other health-related variables (insurance, disease types, health conditions, etc.) were among the most frequently used variables in these models. Among the health-related issues, tuberculosis, and the prevalence of infectious diseases among homeless people was the most researched topic (n= 6). Substance abuse and other drug-related epidemic (n=3), access to basic health care services (n=3), emergency department visit (n=2), and other community issues (n=5) also came upon in the reviewed studies.

### 3.2 Data source and types of geospatial analysis

Among the reviewed articles, ArcGIS was the most popular geospatial analysis software used by the researchers for the health research of homelessness. Additionally, studies used R statistical Package, R-INLA, Geo-Mapping Software (GMS), WebGIS, and STATA for both statistical and spatial analysis. These studies used the aforementioned geospatial analysis software and tools for data visualization and mapping ^11,21-23^, distribution of homeless center ^24-26^ or visualize results ^19^ related to the health-related problems and homelessness.

Several studies used point density mapping ^18,27^ or Kernel density estimations to analyze the pattern of transmission of diseases, injuries, drug use, etc. ^20,22,24,28,29^. Hotspot and cold spot analysis in GIS is another frequently used geospatial tool for mapping the patient’s location of different diseases ^18,30^, discarded needles ^9^, and homeless women veteran ^22^. Some studies also employed buffer analysis or proximity analysis to comprehend the accessibility pattern or activity space of currently homeless or previously homeless peoples ^4,5,22,31^, or applied proximity analysis in ArcGIS to identify the spatial access to family planning clinics for a homeless woman. Along with all these aforementioned widely used geospatial analysis techniques, researchers also used choropleth map, cluster analysis ^9^, and overlay analysis ^12^ for exploring the nexus between homelessness and public health issues.

Most of the studies included in this review reported using data from official federal or governmental data source such as the Centers for Disease Control and Prevention (CDC) in the USA; Regional Public Health Department of Montreal in Canada; Office for National Statistics and Health and Social Care Information Centre (HSCIC) in the UK; Institute for Clinical Evaluative Sciences, Canada; All-Payer Claims Databases; Statewide Planning and Research Cooperative System (SPARCS); New York State Department of Health; London Tuberculosis Register; Westminster City Council; Department for Work and Pensions (DWP), UK; Department for Education (DoE), UK. Very few studies used primary survey data ^4,11,21,28,31,32^, focus group discussion data ^19^, key informant interview data ^24^ for the health-related studies of the homeless.

The analysis of data sources reveals an important finding that the geolocation data of homeless people and patients of different diseases are important in the research of health issues of the homeless. In general, developed countries collect and preserve the geolocation data of the patients and homeless peoples. Therefore, all the studies related to the geospatial analysis of homelessness and public health are from these countries. No research studies on the public health of homeless people using geospatial analysis are published from the developing or less developed countries, which raises a major concern regarding equity and equitable distribution of resources and lack of research in the underdeveloped countries for those who are in need the most.

### 3.3 Homelessness and health: Geospatial findings from the review

Homelessness is a major social and public health crisis that is being increased over time and associated with dire implications for public health ^33^. Even though the application of geospatial analysis techniques is comparatively low in number, the history of this homelessness-public health concern-spatial analysis nexus has a historical root. In this study, we focused specifically on identifying the key characteristics of the geospatial tools and techniques used in homeless research with a public health focus, the types of data used in these studies, examined the problems and prospects in doing this. We also stretched our focus towards the issues these studies focused on-directly or indirectly related to public health.

#### Geospatial techniques used in health research among the homeless population

##### Homelessness and diseases (contagious and noncontagious) pattern

Previous studies used spatial analysis techniques to understand the patterns, trends, and relationships of different contagious and noncontagious diseases with homelessness. For example, several studies used spatial analysis tools to examine the patterns of tuberculosis transmission through GIS-based mapping and visualizations. This mapping and use of geospatial analyses provided information on whether there is a significant relationship between homelessness and disease transmission patterns ^18,24,29,30,32^.

These studies collected geographic locations of the tuberculosis patients and visualized the pattern of disease through ArcGIS or other GIS-based mapping tools. These studies also used point density analysis and hotspot analysis to identify spatial clustering of infected patients with these contagious diseases. Findings from these studies indicated that people who are currently homeless or been homeless at some stages of their lives are more susceptible to infectious diseases.

Other infectious diseases such as Methicillin-resistant Staphylococcus aureus (MRSA) infection or blood-borne and sexually transmitted diseases were also mapped and visualized in previous studies that seek to explore the infection pattern and examine the association between infectious diseases and homelessness ^23,28^. Even though these studies used both statistical and spatial analysis techniques as their analytical tools, these mapping and visualizations aided these studies to both locate the homeless people and visualize whether these locations represent an association with the diseases.

Some studies also reported a significant relationship between homelessness and other noncontagious diseases, and they utilized different geospatial analytical techniques. For example, using hotspot analysis in ArcGIS, Goswami et al. (2012) identified that 40% of the HIV-infected patients had lived in a homeless shelter. The same study utilized georeferencing techniques to identify the location of the homeless shelters before using the kernel density tool to identify the hotspots ^32^. Similarly, other studies found that homeless centers report a higher number of patients with psychiatric disabilities ^21^. In most cases, these studies utilized kernel density tools in ArcGIS to perform hotspot analysis and identify spatial patterns of diseases spread and the association between patient locations and locations of homeless shelters/centers.

##### Homelessness and other health-related issues

Even though the majority of the reviewed studies focused on examining the trends and patterns of contagious and noncontagious diseases among homeless people, few of the studies extended their efforts to understand some ongoing epidemiological problems-i.e., opioid overdose, substance abuse, etc. These studies investigated the data on opioid overdose data, geospatial distribution of discarded needles in the city ^9^. For that purpose, studies utilized choropleth maps, cluster analysis using Local Moran’s I, and hotspot analysis. The spatial autocorrelation analysis reported the high discarded needle density among homeless populations and homeless centers.

Similar to the studies that examined contagious and noncontagious diseases, these studies focused on epidemics also used spatial analysis tools for visualization and mapping and hotspot analysis to identify the clusters, patterns, and associations between homeless populations and centers and the location. All these studies reported a higher correlation between the homeless population and their location and the health issue of interest. From the opioid overdose epidemic, discarded needles, and the location of the people who inject drugs (PWID), all these issues are highly found among the people who are currently homeless or were homeless at some point in their lives ^9,27^.

Additionally, some studies utilized spatial analysis techniques to explore the post-disaster health crises among the people who became homeless as a result of a disaster (Curtis et al., 2015; Doran et at., 2016; Lee et al., 2016). Doran et al. 2016, examined the emergency department (ED) visits among the homeless population in the weeks after Hurricane Sandy’s landfall. They utilized both spatial and statistical analysis tools to visualize and calculate the association between the events. The study reported a significant increase in ED visits after Hurricane Sandy. Similarly, Lee (2016) also reported that emergency department visits increased among the patients who were elderly, homeless, or drug dependent. Both these studies used the geographic distribution of post-hurricane Sandy emergency zone visits and utilized spatial analysis techniques to map and visualize the distribution of the same.

##### Homelessness and social issues

Even though the articles from this review focused primarily on the health problems of the homeless population, they also emphasized some other social issues that are almost as important as the health-related problems. These issues include the accessibility of the basic services ^4,22^, community integration ^4,5^, the pattern of injuries ^20^, gambling ^12^, neighborhood distress ^11^, and shelter environment ^19^. Even though the spatial analysis tools used in these studies are very similar to those used in the disease-related studies, the focus and outcome are different. For example, the study by Gawron et al. (2017) used the hotspot analysis, cluster analysis, and spatial autocorrelation analysis using Moran’s I to identify the spatial access to family planning clinics for homeless women.

Another study by Chan et al. (2014) examined community integration along with access to services adopting a similar approach. This is the only study in our review that incorporated participatory mapping techniques alongside the traditional computer-aided GIS platforms. Findings from this study reported that community integration does not differ significantly based on the demographic characteristics or housing types. Other studies used density maps and kernel density analysis to identify the spatial patterns and distribution of injuries ^20^. On the other hand, similar studies reported a higher correlation between gambling, shelter environment, and neighborhood distress with the location of the homeless shelters ^11,12,19^.

**Table 1.** Summary of the reviewed studies

| Study | Country | Spatial analysis techniques | Data used | Data source | Tools | Key findings |
| --- | --- | --- | --- | --- | --- | --- |
| Agarwal, Nguyen <sup>18</sup> | USA | Point density analysis and mapping<br>Hot spot analysis | Case locations, Time of infection, genotype, social and demographic characteristic of the patients | CDC | ArcGIS | 5 common GENType clusters seen from 2009 to 2015<br>Spatio-temporal distribution of GENType was significant and was correlated with ethnicity, alcohol use, homelessness etc. |
| Aho, Lacroix <sup>30</sup> | Canada | Hotspot analysis of the risk factors<br>Spatial analysis of the patient's locations | Intravenous drugs, tobacco and alcohol use, sex work, mental health disorders, Hepatitis-C and HIV coinfection, homelessness, prior incarceration records | Regional public health department | Not Mentioned | Substance use among the marginalized found to be influencing infection rate<br>Alcohol and substance use in addition to homelessness was reported as most severe factors causing infections |
| Tosas Auguet, Betley <sup>23</sup> | UK | Maps for significant predictors of MRSA infection | Population density, health characteristics, housing characteristics, demographic variables | ONS, UK; HSCIC, UK | R-INLA built-in model | High risk of CA-MRSA is linked with overcrowding, homelessness, low income, and recent immigration<br>Spatial distribution of areas is highly correlated with exposure risks |
| Bearnot, Pearson <sup>9</sup> | USA | Choropleth map, cluster analysis with Local Moran's I to determine statistically significant hot spots, cold spots, and outlier clusters. | 311 service requests data; tweeter data for needle cleanup | Open Data Commons Public Domain Dedication and License (PDDL) | R statistical Package, ArcGIS | More than two-thirds of the discarded needles were found within 1-km of safe deposit sites and homeless shelters<br>Hotspot analysis using GIS techniques are useful tool to leverage publicly available data for better decision-making |
| Brusilovskiy and Salzer <sup>21</sup> | USA | Data visualization<br>Hotspot analysis using choropleth maps | Economic, social, community, demographic | Primary survey; ACS | GIS | Community-level indicators are marginally correlated with well-being<br>sociodemographic characteristics reported <u>higher correlation with wellbeing factors</u> |
| Callahan, Hwang <sup>34</sup> | USA | Hotspot analysis<br>Spatial interpolation | Recovery homes data, homelessness, unemployment data, substance use | Oxford House Illinois Inc. | ArcGIS | Recovery houses were often located in areas with higher unemployment rate.<br>Location of the houses create inequality in access to citizen services |
| Chan, Gopal <sup>4</sup> | USA | Euclidean distance analysis, multi-ring buffer analysis and spatial weighted matrix development | Location of previously homeless people, Location of basic services | Primary survey; MassGIS data layer | ArcGIS | Activity spaces reported to be inversely correlated with available community features<br>Proximity to health care services influences activity spaces and spatial distribution of communities |
| Chan, Helfrich <sup>5</sup> | USA | Activity spaces were created in GIS from the locations identified on participant map | Location of previously homeless people | Primary survey; MassGIS data layer | ArcGIS | No significant relationship between activity space size and community integration measures |
|  |  |  |  |  |  | Despite the positive relation between use of homeless services and activity spaces, it negatively affects other social and psychological integration efforts. |
| Curtis, Curtis <sup>24</sup> | USA | Mapping homeless and Kernel density estimations (KDE) of all mentions of SVG | Location data, post-disaster recovery, crime, mosquito control and tuberculosis data of the neighborhood | Interview; video survey data from car | GIS | SVG could provide a viable alternative in places where spatial data are non-existent.<br>Lifestyle of homeless population could be mapped using this proposed technique |
| Curtis, Felix <sup>10</sup> | USA | Kernel Density Estimation to identify spatial clustering of overdoses location | themes and key words associated with drug activity, homeless camp location | Los Angeles County Public Health Department, Primary Survey | ArcGIS, google Earth | Opioid overdoses are strongly connected with homelessness camp locations<br>Structural violence is traced in some solutions proposed and implemented among the homeless peoples |
| Cusimano, Chipman <sup>20</sup> | Canada | Density napping of patients of fall injuries, mean household income and homeless center using kernel density function | Demographic and land use variables- such as household income, age, and the location of homeless shelters | Institute for Clinical Evaluative Sciences, Canada | ArcGIS; web-based GIS | Homeless shelter's location was reported as one of the explanatory factors for the spatial distribution on injuries<br>People living in low-income areas are shown to be at the highest risk of injury |
| Dickson-Gomez, McAuliffe <sup>11</sup> | USA | Spatial join of survey data to census track to measure distress value | Location and living pattern of low-income residents or homeless people | Primary survey; American Community Survey (ACS) | ArcGIS | Significant mismatch reported between housing choice among those with housing subsidies (more) and those in supportive housing perceived (less).<br>No relationship was found between low-income residents with housing subsidies and living in high stress neighborhood |
| Doran, McCormack <sup>25</sup> | USA | Mapping, spatial analysis and z score calculation of ED visits for homelessness | Demographic characteristics, insurance status, geographic distribution, and health conditions | All-Payer Claims Databases | Stata, ArcGIS | Statistically significant increases in ED visits for diagnosis codes of homelessness or inadequate housing in the week after Hurricane Sandy's landfall.<br>Secondary diagnoses among the same group also differed after versus before Hurricane Sandy |
| Doran, Johns <sup>35</sup> | USA | Linked data analysis | Demographic characteristics, insurance status, geographic distribution, and health conditions, emergency shelter data | NYC Department of Homeless Services' CARES database, | SAS PROC LIFETEST | 5% of the patients in the sample who were not homeless at their ED visit entered a shelter within 1 year<br>Majority of these group entered the shelter within 1 month after their ED visit |
| Gawron, Pettey <sup>22</sup> | USA | Data visualization, proximity analysis, hot spot analysis | Sociodemographic, population density, travel behavior, religion, ethnicity | American Community Survey (ACS) | ArcGIS | Homeless women veterans represent the most vulnerable subset at greatest risk for adverse reproductive outcomes.<br>Distance to VAMC is significantly higher compared to distance to VHA facilities. |
| Goswami, Hecker <sup>32</sup> | USA | Geocoded patients address and create density maps using the kernel density tool | Location and disease type data of patients | Primary survey | ArcMap | More than one-third (40%) of the patients had lived in a homeless shelter<br>GIS-based screening could be effective in identifying populations with high disease burden and poor healthcare access |
| Horan and Van Hout <sup>27</sup> | Ireland | GPS mapping and visualization of the patients' locations | Location of homeless center, PWID data, supervised injecting facilities (SIF) data, locations of overdoses | Health Protection Surveillance Centre | Geo- Mapping Software (GMS) | 43.1% of total PWID sample are currently homeless<br>Significant correlation was found between history of accidental overdose, injecting alone and previous history of overdose |
| Lee, Smith <sup>26</sup> | USA | Mapping geographic distribution of post-Hurricane Sandy emergency zones of New York City. | Evacuation zone location, patient's data, demographic data, Insurance type | SPARCS, New York State health Department | Stata, ArcGIS | Patients who were elderly or homeless, or drug dependence were more likely to visit emergency departments after Hurricane Sandy<br>Statistically significant decreases in overall post-Sandy emergency department use but increased utilization in the most vulnerable evacuation zone |
| Logan, Jolly <sup>28</sup> | Canada | Network graphs to map high frequency geographic areas<br>Kernel density estimation (KDE) to infer high risk areas and cluster | Injection drug users (IDU) social network data, street youth, men who have sex with men, and homeless people | Primary survey | ArcGIS | The social network of homeless people is connected to disease risk space |
| Pina, Alves <sup>36</sup> | Brazil | Kernel density estimation<br>Getis-Ord Gi* technique | Sociodemographic data, social vulnerability (low income, poor housing conditions and homelessness), occurrence of pneumonia cases | Demographic Census of the Brazilian Institute of Geography and Statistics | ArcGIS | Cases are concentrated in regions with greater social vulnerability (low income, poor housing conditions and homelessness)<br>Unequal access to health care facilities based on the location of the patients |
| Richards and Smith <sup>19</sup> | USA | Mapping of food resources source using GIS | Food resources, food prices, Food access and availability, income, education, no. of children per household, shelter data | Focus Group Discussion (FGD) | ArcGIS | The shelter environment and surrounding community influenced lifestyle factors related to health, including food access and availability, exercise behaviors, job access, and day care issues. |
| Smith, Trienekens <sup>29</sup> | England | Used kernel-density estimation and k-function analysis to assess spatiotemporal progression and geographic clustering of the outbreak | Drug use, prison link, homelessness, alcohol dependence, sex, and ethnicity | London Tuberculosis Register (LTBR), Enhanced Tuberculosis Surveillance System | R, Stata | Homelessness is a major social risk factor which is responsible for the outbreak in 25% cases.<br>Treatment time varies based on patients' association with risk factors- lower for those with higher risk |
| Townley, Pearson <sup>31</sup> | USA | Mapping of activity spaces and proximity analysis in GIS | Sense of community, psychological well being | Primary survey | ArcGIS | Youth engagement with service-related activities is highly correlated with sense of community and psychological wellbeing.<br>Homeless youth's have significant involvement in community activity spaces. |
| Wardle, Asbury <sup>12</sup> | UK | Weighted spatial overlay analysis | Substance use, gambling, financial debt, unemployment, ethnicity, mental health, homelessness | ONS, HSCIC, DWP, City council | ArcGIS | Gambling is associated with the homelessness shelter's locations<br>Risk factors varies with the spatial distribution of high-risk locations |

## 4. Discussion

### 4.1. Geospatial analyses in homelessness and public health research

Spatial analysis and different geospatial analytical tools and techniques have been used in health research since the 1960s ^13,14^. The advancement in these spatial analysis techniques paved a robust pathway to examine many complex health-related issues emphasizing on more specific target groups. The homelessness phenomenon is a global crisis and with the ongoing pandemic, it is crucial now more than ever. Studies already pointed out that unequal access to housing has an association to the infection rates in the USA ^1^. Even before the pandemic, the homelessness problem was prevailing, and these are the same group of people that usually are counterproductive to any social, psychological, and public health improvement integration efforts ^4,5^. The use of geospatial analysis is not only beneficial in identifying the location of these populations, but also could benefit the scientific community and policy makers with detailed insight on the problem and aid in preparing more comprehensive plans. Perhaps that is why, though limited in numbers, there are attempts to study homelessness– public health issues– geospatial analysis nexus.

In this study, we reviewed, synthesized, and offer a critical understanding of how past studies examined the use of geospatial analysis in health-related issues among the homeless population. The findings indicated that these studies used spatial analysis to examine the disease pattern and social issues among the homeless. However, these two aspects are overlapping and has a major influence on each other and highly correlated with the state of being homeless. It is apparent from the findings that the relationship between GIS and health research among homelessness issues is continuously evolving at a global scale. The pragmatic aspect of spatial analysis and rapid development of GIS technologies has been contributing health researchers in understanding the connection between poverty, community integration, the transmission of diseases, and homelessness.

The findings from this review showed that the majority of the health-related studies concerning homelessness focused on the pattern and trend of both contagious and noncontagious diseases ^18,21,24,28,32^. These studies used geospatial analysis techniques to identify the location of the homeless population and shelters, whether there is any clustering of diseases. These same studies also used different statistical modeling approaches to identify the correlation between different variables and homelessness. Beyond the mapping and visualization, hotspot analysis using the kernel density functions is the most prominently used spatial analysis techniques in the reviewed articles ^9,10,18,22,24,28-30,34,36^. However, a few studies also used global and local Moran’s I to identify the spatial autocorrelation between the locations and distribution ^9,12^. In addition to the contagious and noncontagious diseases, some studies also examined issues with epidemiological issues such as opioid overdose and needles discard rate in and around the homeless shelters ^9,10^. These studies also used hotspot and cold spot analysis to identify whether there is any correlation between the location of the homeless shelters and the location of reported opioid cases and needle discard density ^10^.

Although low in number, some studies also examined the relation between some social issues i.e., gambling problems, neighborhood environment and homelessness using spatial analysis techniques ^11,12^. Similar to the studies mentioned earlier, these studies also examined spatial autocorrelation, cluster analysis and geolocating the patients and homeless shelter’s location for mapping and visualization purposes using GIS. Even though ArcGIS is the most used GIS application, studies also used statistical applications with spatial analysis capabilities (i.e., R-INLA, SAS, STATA). Most of these studies used secondary data from federal and regional service provider (i.e, CDC), but few studies (n=5) used primary data collection methods for their studies-focus group discussion (n=1), key informant interview (n=1) and survey methods (n=3).

One of the most striking findings from the review is the geographic locations of the case study sites. All these studies are from the developed countries in the global north. These are the countries that have a more developed health care system where they collect and store location data of the health care service recipients. The same is applicable for the location of the service providers, reported incidents and other geographical data. As these data are available either publicly or upon request, carrying out such research is more accessible in these countries. Most of these studies include data from Center for Diseases Control (CDC) for contagious and noncontagious disease related data (Agarwal et al., 2019), American Community Survey (ACS) for demographic data in the US (Dickson-Gomez et al. 2016; Gawron et al., 2017; Brusilovskiy and Salzer, 2012), and similar national and regional statistical and health service data providers in other countries as well (Tosas Auguet et al., 2016; Aho et al., 2017). Even though most of the studies used secondary data, studies based on primary data were also available (Chan, Gopal et al., 2014; Curtis et al., 2015; Gowsami et al., 2012; Logan et al., 2016; Richards and Smith 20106; Townley et al., 2016). These studies are more easily replicable in the less developed and developing countries. In reality, these less-developed countries will provide additional insight on types of problems someone may face in using such approaches. As it is evident from past studies that these countries are facing rapid urbanization and urban population growth ^37,38^, informal housing and lack of formal housing provisions ^39,40^, lack of coordination and management among public service institutions in all services ^41^ and most importantly, may not have adequate financial resources ^42^. Additionally, as mentioned above, despite the potential to use the spatial analysis techniques in more diverse ways, most studies used it only to visualize and map the locations. Though the mapping is beneficial, the advancement in geospatial technologies could be beneficial for more rigorous analysis of the same problems.

Several studies in this review used cluster analysis that could be extended to temporal analysis to identify the trend of clusters, locations of the same issues in the past and track the trend in the future. Additionally, none of the studies extended their efforts to spatially integrate the factors behind homelessness and health-issues with other socio-economic and environmental factors. It is essential to identify the causal factors, their spatial variations as well as the correlation among them. That would lead to addressing the health problems among homeless people more comprehensively rather than as an isolated issue. Even though studies used the fixed locations of the homeless shelter and centers, advancements in the spatial analysis tools and techniques now allow to track near-real time movement of the people. One of the most important characteristics of the homeless population is that their location may not be as fixed as the other parts of the population. Using geospatial technologies would help to identify the issue and track the potential to move beyond that location. This feature of GIS has been extensively used during the coronavirus pandemic. Several countries used the geolocation feature to track the transmission pattern. Additionally, GIS was used to track transmission and case distribution patterns spatially among different locations both within and across countries. It aided the state to develop more robust lockdown and prevention plans and to prepare themselves for the coming outbreak instead of reacting once it occurs. Similarly, these recent features of GIS could aid in homelessness research. Prior studies used the tool to identify the location of the contagious and noncontagious diseases as well as epidemiological issues. GIS could further help in restricting these diseases, limiting the exposure to the factors causing these issues in the first place and also track the population that needs immediate and long-term services on these issues.

### 4.2. Limitations and future research opportunities

Even though our scoping review reveals the broad application of geospatial analyses to conceptualizing health problems and homelessness issues, it has a few limitations. First, our selection criteria may have missed articles related to GIS or spatial analysis, public health, and homelessness, as GIS terminology is still evolving. Second, we only included peer-reviewed publications that used geospatial analyses techniques to investigate any health or health-related problems among homeless people. We excluded preprints, conference proceedings that did not go through the peer-review process which can offer more critical insight on the subject matter. Third, neither we aimed to assess the quality, risk of bias, or methodological rigor of the published articles, nor we conducted a meta-analysis. We recommend that future studies conduct a meta-analysis on specific GIS and geospatial techniques-related topics and evaluate the risk of bias and quality of the methodology used in the scientific literature.

## 5. Conclusion

This study systematically reviewed the use of geospatial analytical methods and techniques in health research among homeless people. The review revealed the key characteristics of the spatial analysis tools and techniques past studies on homelessness used. Even though the usefulness of geospatial techniques in health-related problems are well-established, the majority of the studies with homelessness so far was restricted in analysis and identification of the trends, patterns, and disease clusters in addition to the mapping and visualizations of the patient locations and clusters. However, there are far more opportunities to use geospatial analysis techniques in homelessness research. Tracking the transmission patterns and distribution of the people as well as tracking their movement, connections to different communities and services and accessibility concerns could be well addressed by integrating spatial analysis into the research. This would aid the policymakers in better preparing to encounter any health-issues among the homeless and develop more realistic policies to encounter the issues. Our review also revealed an important finding that the geolocation data of homeless people and patients of different diseases are important in the research of health issues of the homeless. In general, developed countries collect and preserve the geolocation data of the patients and homeless peoples. That is why, most of the studies in our review are from three countries from the global north. No research studies on the public health of homeless people using geospatial analysis are published from the developing or less developed countries, which raises a major concern regarding equity and equitable distribution of resources and lack of research in the underdeveloped countries for those who are in need the most.

## Data Availability

All data produced in the present work are contained in the manuscript

